# Using Social Media to Help Understand Long COVID Patient Reported Health Outcomes: A Natural Language Processing Approach

**DOI:** 10.1101/2022.12.14.22283419

**Authors:** Elham Dolatabadi, Diana Moyano, Michael Bales, Sofija Spasojevic, Rohan Bhambhoria, Junaid Bhatti, Shyamolima Debnath, Nicholas Hoell, Xin Li, Celine Leng, Sasha Nanda, Jad Saab, Esmat Sahak, Fanny Sie, Sara Uppal, Nirma Khatri Vadlamudi, Antoaneta Vladimirova, Artur Yakimovich, Xiaoxue Yang, Sedef Akinli Kocak, Angela M. Cheung

## Abstract

**Background:** There remains significant uncertainty in the definition of the long COVID disease, its expected clinical course, and its impact on daily functioning. Social media platforms can generate valuable insights into patient-reported health outcomes as the content is produced at high resolution by patients and caregivers, representing experiences that may be unavailable to most clinicians.

**Objective:** We aim to determine the validity and effectiveness of advanced NLP approaches built to derive insight into Long COVID-related patient-reported health outcomes from social media platforms.

**Methodology:** We use Transformer-based BERT models to extract and normalize long COVID Symptoms and Conditions (SyCo) from English posts on Twitter and Reddit. Furthermore, we estimate the occurrence and co-occurrence of SyCo terms at any point or across time and locations. Finally, we compare the extracted health outcomes with human annotations and highly utilized clinical outcomes grounded in the medical literature.

**Result:** Based on our findings, the top three most commonly occurring groups of long COVID symptoms are systemic (such as “fatigue”), neuropsychiatric (such as “anxiety” and “brain fog”), and respiratory (such as “shortness of breath”). Regarding the co-occurring symptoms, the pair of ‘fatigue & headaches’ is most common. In addition, we show that other conditions, such as infection, hair loss, and weight loss, as well as mentions of other diseases, such as flu, cancer, or Lyme disease, are among the top reported terms by social media users.

**Conclusion:** The outcome of our social media-derived pipeline is comparable with the outcomes of peer-reviewed articles relevant to long COVID symptoms. Overall, this study provides unique insights into patient-reported health outcomes from long COVID and valuable information about the patient’s journey that can help healthcare providers anticipate future needs.

## INTRODUCTION

Post-acute sequelae of SARS-CoV-2 (PASC) or long COVID, broadly defined as delayed recovery from infection with SARS-CoV-2, can occur following severe, mild, or even asymptomatic SARS-CoV-2 infection [1]. Patients with long COVID experience lingering or episodic symptoms for greater than 12 weeks or three months after acute infection [1], [2]. Despite a growing interest in characterizing clinical manifestations of long COVID, no standard framework has been established [3], [4]. Symptoms of long COVID are extremely heterogeneous, and their assessment varies widely among studies.

Multiple pioneering efforts have investigated symptoms of long COVID for hospitalized individuals, representing the minority of people with COVID-19 [5]–[7]. An extensive patient-led survey has been conducted in an outpatient setting to explore the symptoms of long COVID over seven months [3]. The survey analyzed numerous symptoms of long COVID from 3,762 confirmed (or suspected) COVID-19 patients and attributed them to 10 organ systems. To our knowledge, this study is one of the most highly cited articles providing insights into Long COVID for researchers and clinicians. However, as the authors have mentioned, one limitation of the study is the existence of a sampling bias toward long COVID patients who participated in the study. Following their suggestion, more outreach with diverse groups of patients is needed to counter sample bias and better characterize the long COVID phenomenon. This shortcoming motivates the proposed social medial approach to improve understanding of long COVID by filling the information gaps in a more diverse patient population.

The rise of social media platforms has provided researchers and public agencies an unprecedented opportunity to gain insight into personal and population health experiences outside traditional healthcare settings [8]–[11]. Global content on social media platforms is consistently expanding as social media users are expected to increase to 4.4 billion individuals by 2025 [12]. With an appropriate data analytics approach, this data has proven useful in generating insights into emerging health conditions, as seen with Ebola [13], Zika virus [14], and foodborne disease [15].

Health language processing with social media has been challenging yet promising in the era of the COVID-19 pandemic. Numerous studies have used Twitter to investigate populations’ health during and after COVID [16]–[19]. Besides COVID, Twitter posts have been considered a valuable resource for studying public health measures, population characteristics, and the evolution of new medical conditions (e.g., [20], [21]). Nonetheless, processing Twitter data with natural language processing (NLP) tools is not straightforward due to the posts’ highly unique structure, style, length, and vocabulary [22]. In addition to Twitter, Reddit has become a widely studied social media platform for public health tracking due to its larger character limit for posts, providing rich contextual information and support for throwaway accounts [23]. In a recent study, researchers used Reddit data to uncover patient-reported health outcomes and patterns of mental health issues in real-time and across vulnerable groups due to COVID [24].

A close examination of the previous social media studies demonstrates that generating useful insights (i.e., clinical symptoms) from social media platforms would need complex and well-designed NLP approaches [25], [26], [15]. Moreover, adopting and integrating these types of patient-generated data into broader health research and services require a comprehensive evaluation of the data and model and interpretability of the generated insights that can improve patient outcomes. This evaluation task is laborious due to the lack of a ‘*Ground Truth*’ from social media data, requiring the pooling of resources from related literature and engaging subject matter experts [27].

In response to the emergence of long COVID, we built an NLP pipeline to facilitate extracting information from user-reported experiences in social media platforms [28]. In this study, we examined the validity and effectiveness of our NLP pipeline to provide insights into patient-reported long COVID-related health outcomes across two popular social media platforms; Twitter and Reddit. To better understand users’ opinions and experiences, we extracted symptoms and conditions (SyCo) and estimated their occurrence frequency. We compared the outputs with human annotations and highly utilized clinical outcomes grounded in the medical literature. We also tracked SyCo terms over time and locations.

## METHODOLOGY

### Data Collection

We used application programming interfaces (APIs) for the two social media platforms. We hashed all usernames and removed URLs through the de-identification process. To further pseudonymize the data, we transformed special characters in the tweets or Reddit posts to lowercase and extracted contractions. The supplementary materials and our previous work describe more details about the data collection and preprocessing steps [28].

For Twitter searches, we chose relevant hashtags (e.g., *#longcovid, #covidlong*, or *#longhauler*) and words included in tweets (e.g., long-hauler, chronic symptoms, long-term effects). Re-tweets, replies, quotes, and nullcast were excluded from the dataset, as well as any tweets that were not in English. Our Twitter dataset’s average character length and word count were 133 and 23, respectively. For the Reddit data, we targeted specific subreddits such as “r/*covidlonghaulers*” that were appropriate to the long COVID topic. In contrast to the short length of tweets (up to 280 characters), Reddit posts can have up to 40,000 characters, and the longest Reddit post in our data set had 17,060 characters.

### Data Pre-processing

#### Self-report Extraction

While individual posts in our corpus may have had various purposes at the time of posting, such as disseminating news and sharing ideas and experiences, for the purpose of this study, we were primarily interested in posts containing self-reported medical symptoms. We used a combination of the regular expression approach (RegEx) and transformer-based BERT classifier to distinguish between posts of self-reports (explaining personal health status) and other posts, including opinions or news reports. The RegEx approach relies on personal pronouns, as shown in Table 1a. The classifier, which we call MNLI classifier, is a Natural Language Inference model - the COVID-Twitter BERT v2, fine-tuned on the MultiNLI dataset [25]. The classifier annotates posts as either self-report (annotated as “1”) or not (annotated as “0”) and poses the candidate labels (e.g., “my experience”) as either “premise” or “hypothesis”. We ran the RegEx model and the combination of RegEx and MNLI classifier (RegEx+MNLI classifier) on the entire Twitter dataset. We saved the scores, with the averaged performances shown in Table 1b. The standalone RegEx filter outperformed the RegEx+MNLI classifier without substantial loss in precision, as reflected by the F1 score (Table 1b). Although the proportion of self-reports is much higher on Reddit, the same self-filtration approach was applied to Reddit posts to keep consistency.

**Table 1.**
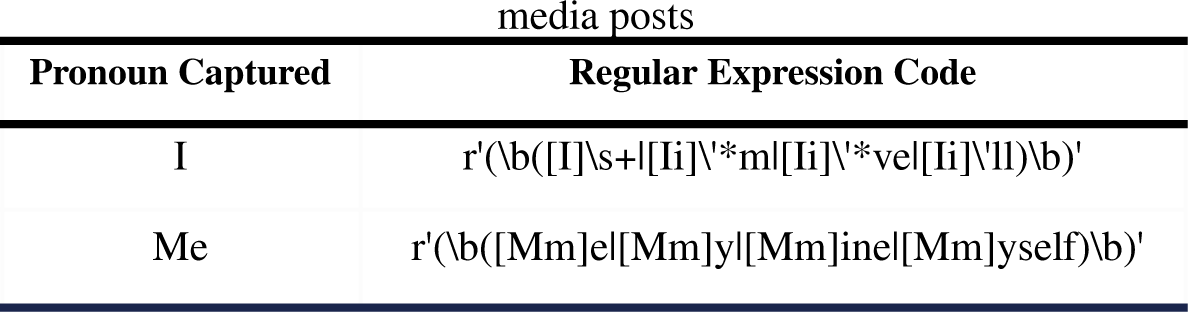
**Table 1a.** Pronouns and Respective Regular Expression (RegEx) used for extraction of self-reports from social

**Table 1b.** The performance results of the self-report filters on the Twitter dataset - RegEx+MNLI classifier is the combination of RegEx, and COVID-Twitter BERT v2 fine-tuned on the MultiNLI dataset.

| Approach | Accuracy | Precision | Recall | F1 |
| --- | --- | --- | --- | --- |
| RegEx+MNLI classifier | **0.83** | **0.79** | 0.71 | 0.74 |
| RegEx | 0.78 | 0.75 | **0.83** | **0.76** |

#### Location Information Inference for Twitter data

For the Twitter dataset, we used the Nominatim API [29] to infer detailed location information such as city, region, and country from user-defined location information attached to tweets. The API uses OpenStreetMap data to make predictions. In cases where user-defined location information could allude to multiple locations (e.g., London is a city in Canada and England), Nominatim API returns the most likely location based on its criteria. For the Reddit data set, location information was not available.

### Extraction and Normalization of Long COVID Symptoms and conditions (*SyCo*)

Transformer-based BERT models were mainly used to extract and normalize long COVID symptoms and conditions, which we refer to as *SyCo* terms in this study. Specifically, we used a UmlsBERT-clinical model fine-tuned on the n2c2 (2010) dataset (Bhambhoria et al., 2022) and for comparison also used the Stanza clinical NER (Stanza-Clinical) from the Stanford NLP group. Given the BERT models’ input length limit, we used rolling windows to segment Reddit posts into smaller chunks with less than 1024 characters.

We implemented a two-step normalization task to analyze our findings, reduce inflectional forms, and compare them with existing works. In the first step, we transformed each extracted raw symptom and condition into its *common base form*. The common base forms for SyCo terms were built in-house through a manual review of all the extracted terms. In the second step, we mapped the symptoms into their corresponding *unique concept* in a target knowledge base (KB) derived from a highly cited and utilized long COVID research article [3]. The KB includes 203 symptoms gathered from 3762 patients with long COVID through an online survey.

The conversion between extracted *raw terms* from social media and either the *common base form* or a *unique concept* in the KB was done using a semantic search approach with BERT bi-encoders [30]. Using this approach, we first created embeddings for all the extracted raw SyCo terms. Then, we retrieved the top *common bases* or *unique concepts* with high semantic overlap with the *raw terms* at the search time. Following a manual review of the retrieved pair, we set a cut-off threshold where pairs with similarity scores greater than the threshold were stored as the match and included in our analysis. For the rest of this paper, *normalized* terms refer to the raw SyCo terms transformed to their common base in the first step of the normalization process. In addition, *mapped* terms refer to normalized SyCo terms further mapped to their corresponding *unique concepts* in the KB.

### Evaluation

To determine how good the BERT models are at extracting the SyCo terms, we created a human-annotated corpus from Twitter and established a proximity-based score to measure potential overlap. Our in-house human-annotated Twitter corpus includes 200 randomly sampled tweets annotated by four trained annotators. The proximity-based score was calculated by dividing the intersection of extracted entities by the union of extracted terms, with duplicated extracted entities removed for annotators and the model. The closer the proximity metric to 0, the closer the model’s predictions to the human-annotated benchmark. Validity and reliability of the extracted and normalized symptoms, a subset of SyCo terms, were evaluated compared with an online survey including 3762 participants from 56 countries [3], [31], [32].

## RESULTS

### Data Overview

Table 2 lists the statistics of the English-language posts collected from Twitter and Reddit. In addition to posts, the timestamp, geographical coordinates (if available), user’s location (user-defined), and user’s profile description were gathered for each tweet. A total of 107 countries were represented in our Twitter sample; most respondents tweeted from the United States (50.7%), followed by the UK (29.9%) and Canada (6.9%).

**Table 2.**
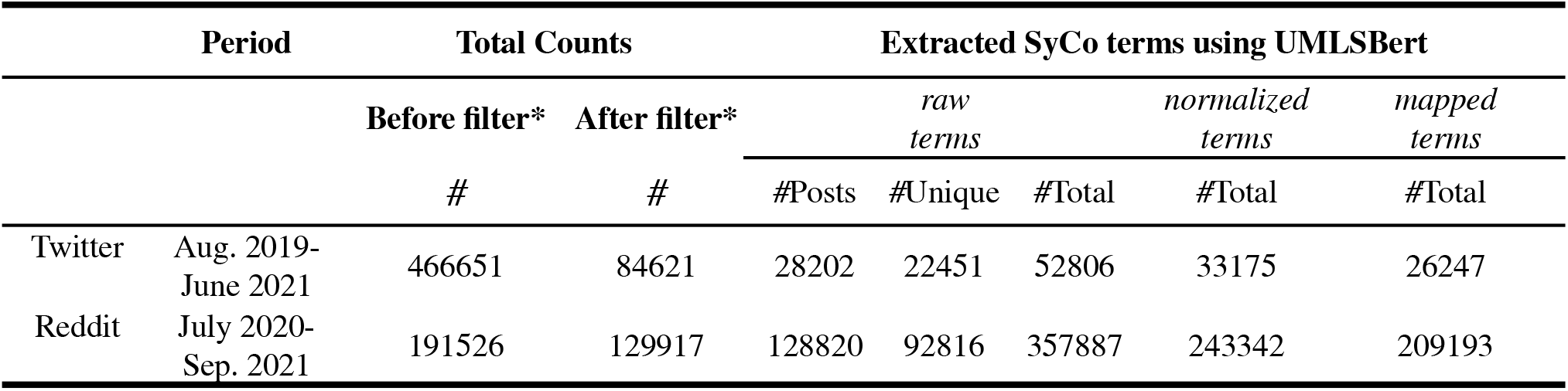
Statistics of Twitter and Reddit data. *The filter refers to the self-report filter. #Posts denotes the count of posts with at least one extracted SyCo term. #Unique denotes the counts of unique extracted SyCo terms. #Total denotes the total occurrence counts of extracted SyCo terms.

### Evaluation of Extraction of SyCo Terms

Table 3 compares the performance of UmlsBERT-Clinical and Stanza-Clinical for entity extraction task from Twitter data. Both models’ performances are compared against human annotators in terms of the proximity score. As shown in the table, we also included entity extraction results by the data augmentation approach UMLS MetaMap (+AMIA) introduced in our earlier work [28]. UMLS MetaMap (+AMIA) uses the MetaMapLite tool to extract entities associating with UMLS’ Concept Unique Identifiers (CUIs) and augments the results with a manually annotated dataset consisting of clinical concepts and colloquial expressions (e.g., brain fog) from tweets. Based on the results, UMLS MetaMap (+AMIA) tends to capture more entities than human annotators; however, some may not be as relevant or provide sufficient insight to experts. Stanza tends to capture fewer entities than human annotators. Consequently, there is the risk of missing information that subject matter experts may consider relevant. UmlsBERT-Clinical has the lowest sum of absolute values for the proximity-based evaluation metric (0.28), indicating predictions closest to those extracted by human annotators from the sample. Hereafter, the rest of the analysis is based on using UmlsBERT-Clinical for the extraction task due to its better overall performance than Stanza.

**Table 3.** Proximity score. The comparison results of the proximity score of human evaluation on 200 tweets were identified by trained annotators with outputs from UmlsBERT-Clinical, Stanza-Clinical, and MetaMap +AMIA. Annotators #3 and #4 have medical backgrounds. The comparison values are based on the proximity-based evaluation metric, defined as the difference between the annotator’s and model’s match counts. The closer the proximity metric to 0, the closer the model’s predictions to the ground truth (human annotator).

|  | UmlsBERT-Clinical<br>[28] | MetaMap +AMIA<br>[28] | Stanza-Clinical |
| --- | --- | --- | --- |
| Annotator 1 | -0.13 | -0.46 | 0.33 |
| Annotator 2 | -0.02 | -0.29 | 0.33 |
| Annotator 3 | 0 | -0.31 | 0.51 |
| Annotator 4 | 0.13 | -0.17 | 0.48 |
| **Sum of the absolute values** | **0.28** | **1.23** | **1.65** |

### Occurrence Frequency Estimation at any point in time

The occurrence frequency of normalized SyCo terms at any point in time is shown in Figure 1. Figure 1a and Figure 1b depict the occurrence frequency of the mapped SyCo terms, and Figure 1c illustrates the occurrence frequency of Normalized SyCo terms. The mapped terms were further categorized by the affected organ systems, similar to the survey study [3], and the aggregated occurrence frequency per each category is shown in Figure 1b.

**Figure 1.**
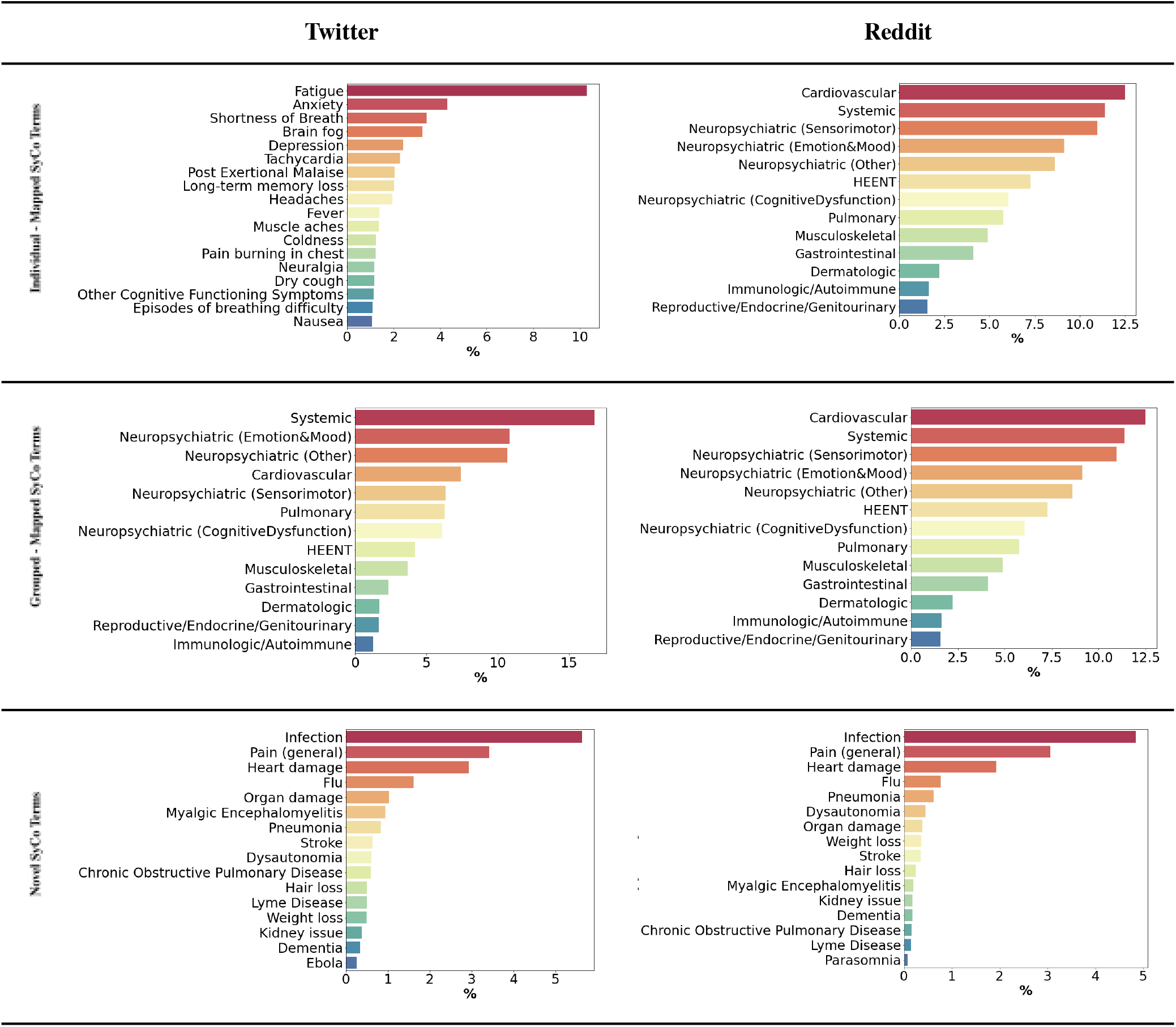
The occurrence frequency of the most prevailing extracted SyCo terms in Twitter and Reddit data with occurrence frequency greater than 1%. Mapped SyCo terms are the nomalized SyCo terms that were mapped to the *unique concepts* in the KB derived from [3]. Mapped SyCo terms were further grouped based on the affected organ system established by [3]; Novel SyCo terms are the unmapped normlized SyCo terms.

#### Comparison of Social Media to the Survey Article

Systemic and neuropsychiatric symptoms were evidenced as the top-occurring symptoms in our study and the survey study. Among these, Fatigue appeared as the most frequent symptom on both Twitter and Reddit. Anxiety (4.8%), Shortness of Breath (3.5%), Brain Fog (3.4%), and Depression (2.7%) were among the top 5 most occurring symptoms on Twitter, whereas for Reddit, they were Anxiety (4.6%), Tachycardia (4.3%), Brain Fog (3.6%) and Shortness of Breath (2.7%). This observation is in line with the survey study [3], where Fatigue, Breathing issues, and Cognitive Dysfunction (e.g., Depression and Anxiety) were reported by patients as the top three most debilitating symptoms. Other terms, including Headaches, Dizziness, Pain, Burning in the Chest, Fever, Nausea, Dry Cough, and Neuralgia, appeared in the top 20 occurring terms in both Twitter and Reddit data; however, with slightly different prevalences (Figure 1a). Immunologic / Autoimmune, Dermatology, and Reproductive / Genitourinary / Endocrine were the lowest three categories of SyCo terms across all three sources - Twitter, Reddit, and Survey study.

#### Discovery of Novel Conditions

Novel SyCo terms shown in Figure 1c are neither reported nor categorized in the survey study [3]. Among novel terms, infection and pain are the top two reported conditions. Other conditions include Flu, Organ Damage, Hair Loss, Weight Loss, Lyme Disease, Dementia, Parasomnia, Pneumonia, Dysautonomia, kidney issues, and Chronic Obstructive Pulmonary Disease (COPD) were among the top 1% of reported terms.

### Co-occurrence Frequency Estimation at any point

Figure 2 shows how often pairs of long COVID-normalized SyCo terms co-occur in both Twitter and Reddit. As expected, the co-occurrence map is more “dense” for the Reddit data (Figure 2a) than the Twitter data (Figure 2b). Since Reddit posts are significantly longer than tweets, they contain more contextual information and repeated SyCo terms. Based on the results, a pair of ‘Fatigue & Headaches’ was among the most co-occurred terms in both platforms. In addition, for Twitter data, the pairs of ‘Fatigue & Shortness of Breath’, ‘Fatigue & Migraines’, ‘Fatigue & General Pain’, ‘Fatigue & Hair loss’, ‘Fatigue & Infection’, ‘Brain Fog & Fatigue’, and ‘Depression & Anxiety’ co-occur more commonly than other terms; for Reddit common SyCo pairs include ‘Fatigue & Bradycardia’, ‘Fatigue & Anxiety’, and ‘Fatigue & Short Term Memory Loss’.

**Figure 2.**
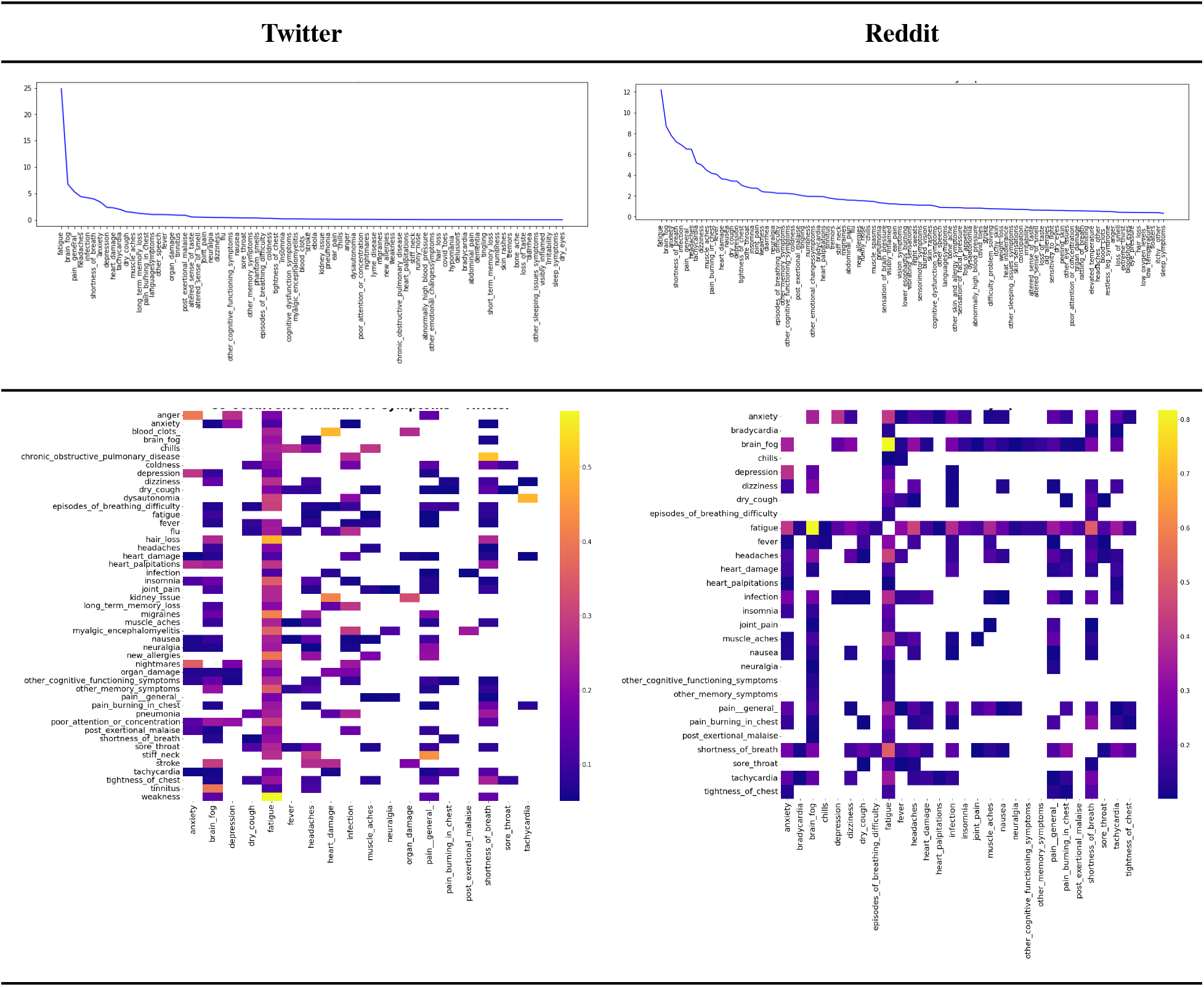
Co-occurrence Frequency of Nomrlized Long-COVID SyCo terms in Twitter (higher than 50%) and Reddit (higher than 10%) data. Higher values are shown by the intensity of pink and blue shading.

### Spatio-temporal Frequency Estimation

The distribution of long COVID-normalized SyCo terms over time is shown in Figure 3 for Twitter and Reddit data. As evidenced by the results, the incidence of the Neuropsychiatric SyCo terms is dominant, followed by the Systemic category, across both social media platforms. On a more granular level, ‘Fatigue’, ‘Anxiety’, and ‘Infections’ were the most prevalent terms reported. Our findings indicate that the predominance of terms varied over time, where for Twitter, ‘anxiety’ was dominant through June 2020, and afterward, ‘Fatigue’ was the most commonly reported symptom. ‘Infection’ has been reported by users as a persistent condition for the entire period. On Reddit, for most periods, ‘Fatigue’ and ‘infection’ were more dominant than ‘anxiety’.

**Figure 3.**
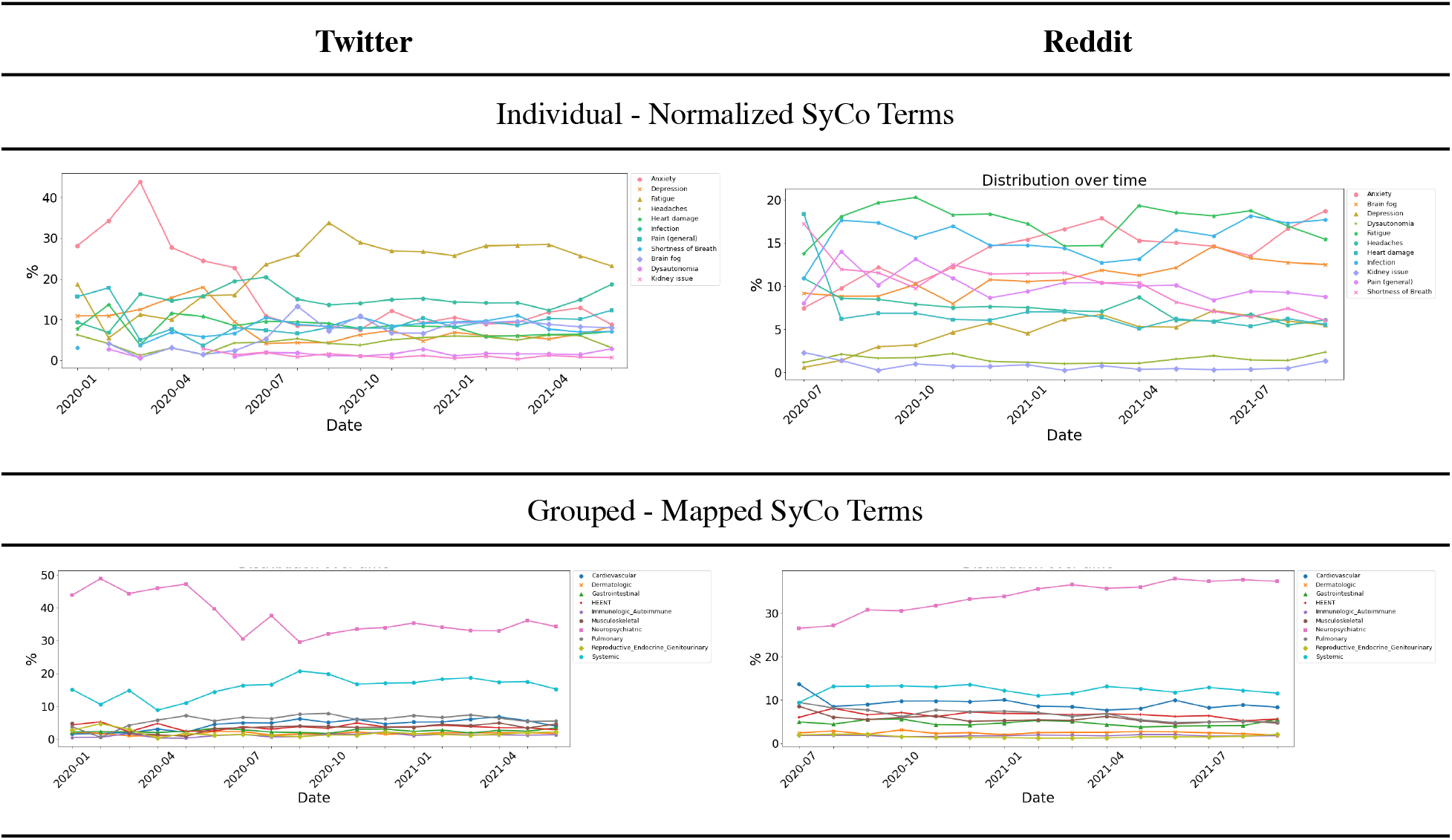
The distribution of long COVID normalized and mapped SyCo terms over time;

Based on our spatial analysis performed on Twitter data, among all the mapped SyCo terms (26247 terms aggregated across all tweets as shown in Table 2), 34% included location information. The 34% were spread across 62 countries, whereby the US (15%), UK (13%), and Canada (2%) were the top three for self-reporting of symptoms related to long COVID (full details are listed in the supplementary materials). The other 59 countries reported fewer than 1% of the SyCo terms and were excluded from our analysis. Figure 4 indicates the proportional contribution of the top four reporting countries to the total occurrence frequency of SyCo terms mapped to *unique concepts in KB* and grouped by the affected organ system.

**Figure 4.**
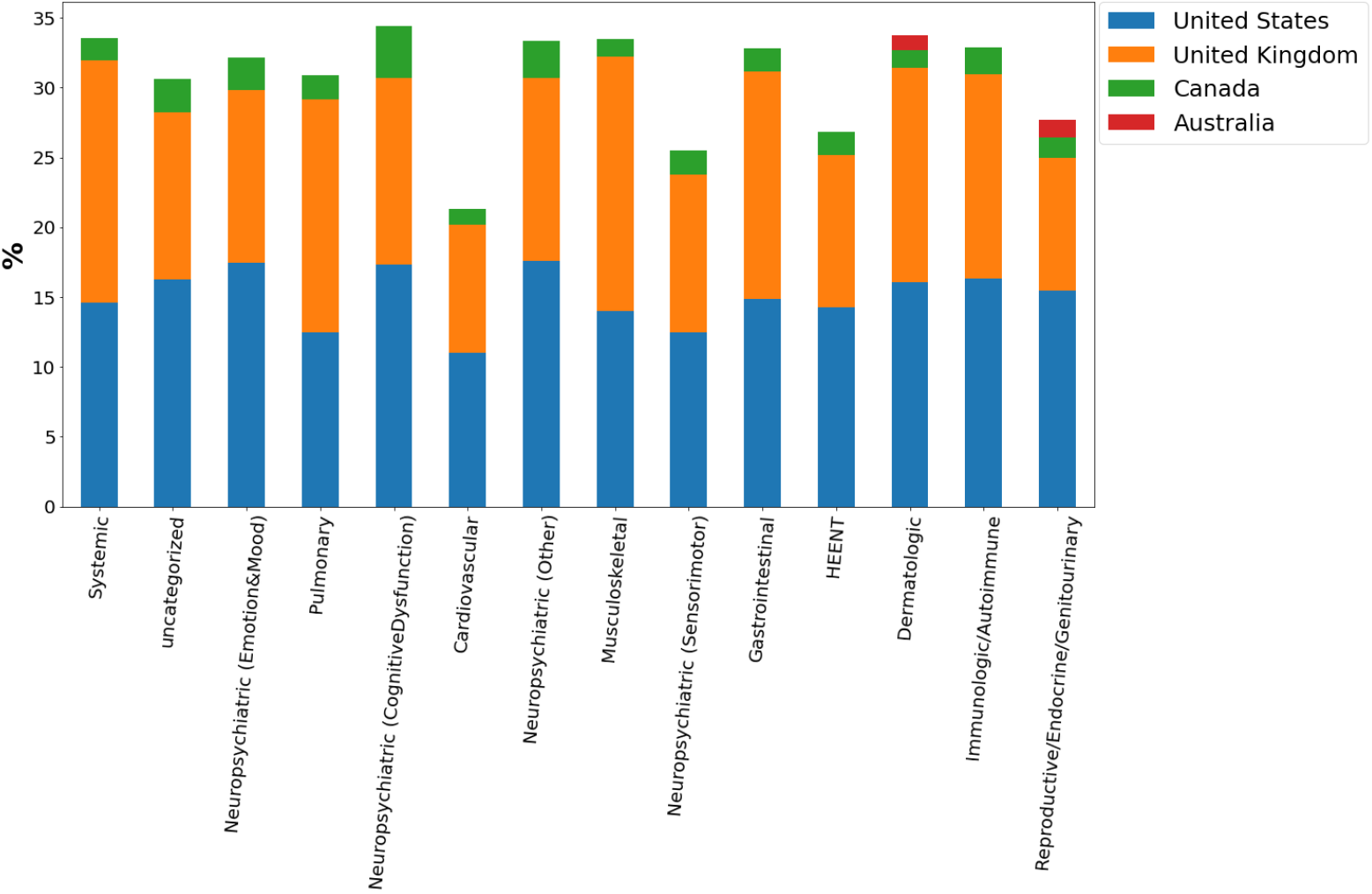
The proportional contribution (%34) of the top four countries (United States, United Kingdom, Canada, and Australia) to each group’s occurrence frequency of SyCo terms. The proportions are measured as a percentage of frequency group-related SyCo terms per each country group divided by the total count of SyCo terms in that group.

## DISCUSSION

The overarching goal of this study was to highlight the possibility of gaining insight into the patient’s experience of long COVID using social media and NLP approaches. User-generated social media data provides a rich yet challenging source of information about the patient journey outside of the healthcare setting. Significant limitations remain concerning recognizing a patient’s lived experience instead of opinion and aligning common vernacular to recognized medical terminology. However, our study has made progress toward narrowing the data quality gap and evaluating the validity and reliability of social media-driven outcomes regarding long COVID using state-of-the-art NLP approaches. Our results suggest that there is value in the learnings and methodologies outlined in this study to gather insights from patient-reported outcomes on social media platforms.

### Principle Results

Our transformer-based entity extraction tool, called UmlsBERT, outperformed Stanza and UMLS MetaMap (+AMIA) for extraction of symptoms and conditions (SyCo) from both Twitter and Reddit. The outcome of our NLP pipeline, normalized and mapped SyCo terms, were comparable with the outcomes of peer-reviewed articles relevant to long COVID symptoms [3], [31], [32]. In particular, there was very high agreement between our findings and those of three different studies investigating long COVID [3], [31], [32]. Our analysis confirms prior findings that COVID-19 infection is associated with long-term effects that are represented by diverse symptoms and conditions affecting multiple organ systems. The top three most commonly occurring groups of long COVID symptoms were Systemic (e.g., “Fatigue”), Neuropsychiatric (e.g., “Anxiety”, “Depression”, and “Brain Fog”), and Pulmonary (e.g., “Shortness of Breath”).

Our findings support the increasing interest in and confirm the need for supplementing clinical observation with user-generated data. In this study, we reported the frequency of co-occurring SyCo terms. Based on our results, the pair of ‘Fatigue & Headaches’ was among the most co-occurred terms in both social media platforms; to the best of our knowledge, expressing a combination of symptoms related to long COVID has been rarely reported in the literature, certainly not from social media based studies. One instance of clinically derived analysis of symptom co-occurrence comes from a US-based retrospective cohort study [32] evaluating long-term symptoms in COVID survivors, where they similarly observe ‘Fatigue’ tends to co-occur frequently with ‘Abnormal Breathing’ or ‘Shortness of Breath’ in patients with long COVID. This analysis’s potential value is connecting SyCo terms early in illness onset with prognostic factors, such as the likelihood of developing long COVID, and the severity or duration of the long COVID condition. In addition, in our study, we extracted long COVID novel terms such as ‘Infection’, ‘Hair Loss’, and ‘Weight Loss’ and reported conditions similar to other diseases such as ‘Flu’, ‘Cancer’, or ‘Lyme Disease’.

One strength of the NLP pipeline developed in this study is its scalability leading to high-level adaptive capacity for other social media platforms or different medical conditions. Our NLP pipeline is well-poised to scale the information extraction process from user-generated data and connect vernacular to recognized medical ontology. In addition, it enables exploring the longitudinal evolution of symptoms, which may be correlated with prevalent SARS-CoV-2 variants at the time of onset to guide insights into variation in disease course associated with different source viral strains.

### Limitations

A common issue with automating entity extraction and normalization is losing the context behind the extracted terms (in the form of independent tokens or words). Without context, it may be difficult to interpret the meaning behind these tokens. For example, the token ‘death’ could be extracted from a personal opinion, an expression of a feeling, or a factual statement; an example of a personal statement is “I think COVID increases the likelihood of death”, whereas an example of a factual statement is “My cousin died of COVID”. In addition, an example of symptom expression tied to the condition is “This long COVID feels like death”. Furthermore, clustering semantically similar extracted symptoms and bringing them closer to predefined standards and common medical terms increases the risk of losing context.

Another limitation of this study relates to our self-report filter. While regular expressions are effective in finding posts containing first-person self-reports, they are also prone to false positives - e.g., mistakenly keeping a post that voices an opinion or reports symptoms on behalf of someone else - and false negatives - e.g., mistakenly discarding a tweet that excludes first-person pronouns but would still qualify as a self-report. We, therefore, considered other context-based approaches, including a fine-tuned BERT classifier. These approaches may reduce false positive and false negative rates; however, further work is needed to annotate sufficient datasets for manually fine-tuning, sweeping, and optimizing hyperparameters.

The nature of social media data and our NLP pipeline introduces bias to our findings which could impact the reliability of our outcomes. Social media content is unlikely to represent the broader population due to demographic biases in the technology uptake, barriers to access, and regional social media platform preferences. Furthermore, only a subset of users are patients who publicly share their experiences with long COVID. Users’ self-reports may also be influenced by prominent opinions reinforced online through news media or other social media ‘influencers’, which add to sample bias. In addition to inherent bias, our analysis is further biased by including only English-language posts.

In social media, data quality is highly variable due to the use of colloquial language (e.g., “I am dying”), brevity or shorthand, grammatical and spelling errors, etc. Tweets were first collected based on the presence of relevant hashtags and keywords (e.g., “long COVID”). While this approach successfully surfaced many relevant tweets, both false positives (e.g., mistakenly keeping a tweet that refers to ‘long haul’ in the context of transportation) and false negatives (e.g., mistakenly discarding a relevant tweet because it lacks or has a typo in, a target hashtag or keyword) can occur. False positives could be reduced by checking for the context surrounding a match, e.g., excluding tweets that refer to long-haul flights. False negatives could be reduced using advanced semantic language models beyond keyword matching, e.g., classifying clinical tweets versus those not.

### Best Practices and Future Directions

In this section, we would like to explain the best practices for researchers and developers interested in applying NLP to social media to facilitate information extraction tasks.

#### Generalizable Models

UmlsBERT is a transformer-based contextual model with better generalizability and reliability than traditional entity extraction models. However, in our experiments, we found that these models may overfit to occurrences of specific tokens such as ‘vid’. As a result, at inference time, incorrect tokens may be captured. A simple solution to extend the capabilities of these models is by looking at the frequencies of captured tokens and devising simple rules to correct the errors. A simple rule-based strategy should also remove variations of the same frequently occurring tokens. Another potential solution for this task would be utilizing symptom ontology and standard normalization procedures to facilitate comparisons between variations of the same token. Literature references and ground truths can also be referred to for this procedure, especially in the case of new illnesses wherein ontologies may not fully capture the experiences that patients are trying to express - for example, ‘Brain Fog’.

#### Longitudinal Analysis

Providing a longitudinal analysis of posts from the same user will enable better characterization of the evolution of symptoms over time. However, it is essential to note that this might pose challenges for ensuring privacy as, for example, the combination of posts may increase the possibility of re-identifying a user. In addition, aggregating their posts may infer illness in patients who have not consented to such an assessment.

## CONCLUSION

In this work, we examined whether the quality of extracted information from social media is sufficient to provide clinical insights for understanding long COVID. We have shown here the applicability of the NLP pipeline to both Twitter and Reddit platforms. The findings of our study support that social media can augment healthcare research by providing insights into diseases that are captured outside usual episodes of clinical care. Moreover, it supports pandemic advance monitoring and response by enhancing the scope of information-feeding risk models. Significant challenges remain in enhancing the accuracy and context with which symptoms are recognized (from vernacular) and interpreted (to medical ontology), which, if resolved, would add to the overall utility of the process. In conclusion, it is early days for using social media in healthcare. While the aggregation of social media-based user-generated data could have benefits at the population level, the evaluations and actionability of data appear limited unless it is used in conjunction with data from established sources.

## Data Availability

Our github repository provides source codes for reproducing the data referred to in the manuscript from Twitter and Reddit data.

https://github.com/VectorInstitute/ProjectLongCovid-NER

## Acknowledgements

The participating companies and medical subject matter experts deserve special credit. Together, over the course of the project Deloitte, Roche, and TELUS provided clinical and machine learning expertise that enabled the pipeline to function. Key contributions by these Vector sponsor companies include original project ideation; review of clinical literature on long COVID; data collection, cleaning, and annotation; filter implementation; normalization; NER modeling; visualization engineering; and interpretation of results. We also thank the contributions of Jennifer Camaradou as a patient advocate for her insights into long COVID and potential benefit from this body of work.

## Notes

### Competing Interest Statement

The authors have declared no competing interest.

### Funding Statement

This study was supported by Vector Institute and its partner companies and medical subject matter experts. The partner companies include Deloitte, Roche, and TELUS.

### Author Declarations

Social media posts from Twitter and Reddit was used in this study. We used Twitter academic API to extract tweets. We hashed all usernames and removed URLs through the de-identification process. To further pseudonymize the data, we transformed special characters in the tweets or Reddit posts to lowercase and extracted contractions.

