## supplementary materials for "Using Social Media to Help Understand Long COVID Patient Reported Health Outcomes: A Natural Language Processing Approach"

### S.1. Twitter Hashtags List

| Long Haulers Specific | General COVID |
|---|---|
| #longcovid | #COVIDTreatments |
| #covidlong | #Covid19 |
| #longcovid19 | #CovidBrainFog |
| #longhauler | #CovidPneumonia |
| #LongHaulers | #CovidInsomnia |
| #covidlonghauler | #CovidLongHauler |
| #StopTheLongHaul | #CovidSucks |
| #LongCovidRecovery | #CovidPhysicalTherapy |
| #LongCovidSymptoms | #CovidVaccine |
| #postcovid19syndrome | |
| #postcovidsyndrome | |
| #CountLongCovid | |
| #PostAcuteCovid19 | |
| #Linger | |
| #Covid | |

### S.2. Number of Tweets per Country List

| Country | Mapped SyCo terms | |
|---|---|---|
| | # counts | % |
| United States | 4850 | 14 |
| United Kingdom | 4316 | 13 |
| Canada | 631 | 2 |
| Australia | 251 | <1 |

| | | |
|---|---|---|
| India | 121 | <1 |
| South Africa | 97 | <1 |
| France | 79 | <1 |
| Germany | 62 | <1 |
| New Zealand | 43 | <1 |
| China | 41 | <1 |
| Albania | 40 | <1 |
| Argentina | 36 | <1 |
| Switzerland | 34 | <1 |
| Netherlands | 31 | <1 |
| Brazil | 21 | <1 |
| Russia | 18 | <1 |
| Italy | 18 | <1 |
| Angola | 17 | <1 |
| Greece | 16 | <1 |
| Spain | 15 | <1 |
| Kenya | 15 | <1 |
| Iran | 14 | <1 |
| Pakistan | 14 | <1 |
| Denmark | 13 | <1 |
| Colombia | 11 | <1 |
| Czechia | 11 | <1 |
| Philippines | 9 | <1 |
| Nigeria | 9 | <1 |
| United Arab Emirates | 9 | <1 |
| Chile | 8 | <1 |
| Malaysia | 8 | <1 |
| Uganda | 7 | <1 |
| Israel | 7 | <1 |

| | | |
|---|---|---|
| Honduras | 6 | <1 |
| Ghana | 5 | <1 |
| Thailand | 4 | <1 |
| Zimbabwe | 4 | <1 |
| Belgium | 4 | <1 |
| Ecuador | 3 | <1 |
| Peru | 3 | <1 |
| Ukraine | 3 | <1 |
| The Gambia | 3 | <1 |
| Poland | 3 | <1 |
| Iraq | 3 | <1 |
| Guyana | 3 | <1 |
| Latvia | 3 | <1 |
| Rwanda | 2 | <1 |
| Bangladesh | 2 | <1 |
| Democratic Republic of the Congo | 2 | <1 |
| Guernsey | 1 | <1 |
| Venezuela | 1 | <1 |
| Austria | 1 | <1 |
| Bahrain | 1 | <1 |
| Indonesia | 1 | <1 |
| Botswana | 1 | <1 |
| Ethiopia | 1 | <1 |
| Costa Rica | 1 | <1 |
| South Sudan | 1 | <1 |
| Mongolia | 1 | <1 |
| Egypt | 1 | <1 |
| Oman | 1 | <1 |
| Mexico | 1 | <1 |

### S.3. Data Creation

GitHub repository: https://github.com/VectorInstitute/ProjectLongCovid-NER
